# Simplifying cardiology research abstracts: assessing ChatGPT’s readability and comprehensibility for non-medical audiences

**DOI:** 10.1101/2025.03.21.25324378

**Authors:** Kabir Malkani, Zachary Falk, Ruina Zhang, Ryan Hughes, Prianca Tawde, Melissa Parker, Griffin P. Collins, Danielle Maizes, Alexander Zhao, Vinay Kini

## Abstract

Artificial Intelligence (AI)-powered chatbots are increasingly utilized in academic medical settings for tasks such as evidence synthesis and manuscript drafting. This study evaluates the ability of ChatGPT, an AI-powered tool, to simplify cardiology research abstracts for non-medical audiences while retaining essential information. A total of 113 abstracts from Circulation were processed by ChatGPT to be rewritten at a 5th-grade reading level. Readability was assessed using word and character counts, Flesch-Kincaid Grade Level (FKGL), and Reading Ease (FKRE) scores, while a panel of five physicians and five laypeople evaluated the simplified texts for accuracy, completeness, and readability. The simplification significantly reduced word and character counts (p<0.0001) and improved readability from a college graduate level to an 8th-9th grade level (p<0.001). Both physicians and laypeople found the simplified abstracts easier to understand, but some patients expressed concerns about oversimplification and missing details. Overall, ChatGPT proved effective in simplifying cardiology research while largely preserving content integrity, though further refinement of AI tools is needed to ensure accuracy.

**Author Summary:** In this study we investigated how artificial intelligence (AI), specifically ChatGPT, can augment comprehensibility of complex cardiology research and thus make it more accessible to people without a medical background. We focused on simplifying abstracts by having ChatGPT rewrite them at a 5th-grade reading level. We analyzed 113 cardiology abstracts from manuscripts published in the journal *Circulation*, measuring readability and word counts before and after the AI simplification process. A group of five physicians and five non-medical participants then reviewed the simplified versions to assess whether they remained accurate, complete, and easy to understand. Our results revealed that ChatGPT significantly shortened the abstracts and made them easier to read, improving readability from a college level to an 8^th^ or 9^th^ grade level. Both medical experts and non-experts agreed the simplified abstracts were clearer. However, some non-medical participants raised concerns that important details might be lost in the simplification process. This highlights a key challenge: while AI tools like ChatGPT can improve access to scientific information, further refinement is needed to balance simplicity with accuracy. Our work underscores the potential of AI in bridging the gap between medical research and public understanding, making complex health information more approachable for everyone.

## Introduction

The integration of Artificial Intelligence (AI) in medicine, particularly through advanced machine learning algorithms and large language models, has revolutionized healthcare by offering innovative solutions to longstanding challenges [1-4]. AI’s applications, such as diagnostic support and personalized treatment planning, demonstrate its broad utility. At the forefront is ChatGPT, a platform that has shown potential to streamline communication, facilitate information dissemination, and enhance decision-making processes [3, 4].

Less well understood is whether large language models could help communicate complex medical information to non-medical professionals such as patients and medical journalists. Technical jargon in medical literature hinders understanding of disease processes and diagnostic/therapeutic options by non-medical professionals. Effective communication of medical information to support informed decision-making remains a significant challenge, particularly among patients who are not medically trained. This study aims to evaluate ChatGPT’s ability to simplify cardiology research abstracts while preserving accuracy and essential details, with the goal of improving comprehension for a non-medical audience.

## Methods

In this study, we analyzed 113 consecutive abstracts and scientific statements published from July 2023 to November 2023 in the journal *Circulation*. Each abstract was processed through ChatGPT-4 with the instruction: ‘Please rewrite the following text to be comprehensible at a 5th-grade reading level, retaining all original information and excluding nothing.’

To assess readability of the original abstracts and the simplified abstracts processed by ChatGPT, we compared word and character counts along with the Flesch-Kincaid Grade Level (FKGL) and Reading Ease (FKRE) scores. FKGL measures text complexity on a U.S. school grade level, while FKRE measures text complexity on a scale from 0 to 100, with higher scores indicating easier readability. We used a paired t-test to compare the mean word and character counts, as well as the FKGL and FKRE scores, between the original and simplified abstracts.

Next, five internal medicine resident physicians evaluated each original and simplified abstract (n=565 person-abstracts) and graded the simplified abstract on 1) accuracy of medical information, 2) completeness of research findings, and 3) a subjective perception of patient comprehensibility on a 5-point Likert Scale. Five non-medically-trained individuals (“laypeople”), all with undergraduate degrees but without graduate degrees, also reviewed and graded the simplified abstracts for readability, acceptability, and ease of understanding using a 5-point Likert Scale.

## Results

The simplification process significantly reduced the word and character counts, from 306 to 259 words and from 2255 to 1535 characters (without spaces), respectively (both p<0.0001; Fig 1A and Fig 1B).

**Fig 1.**
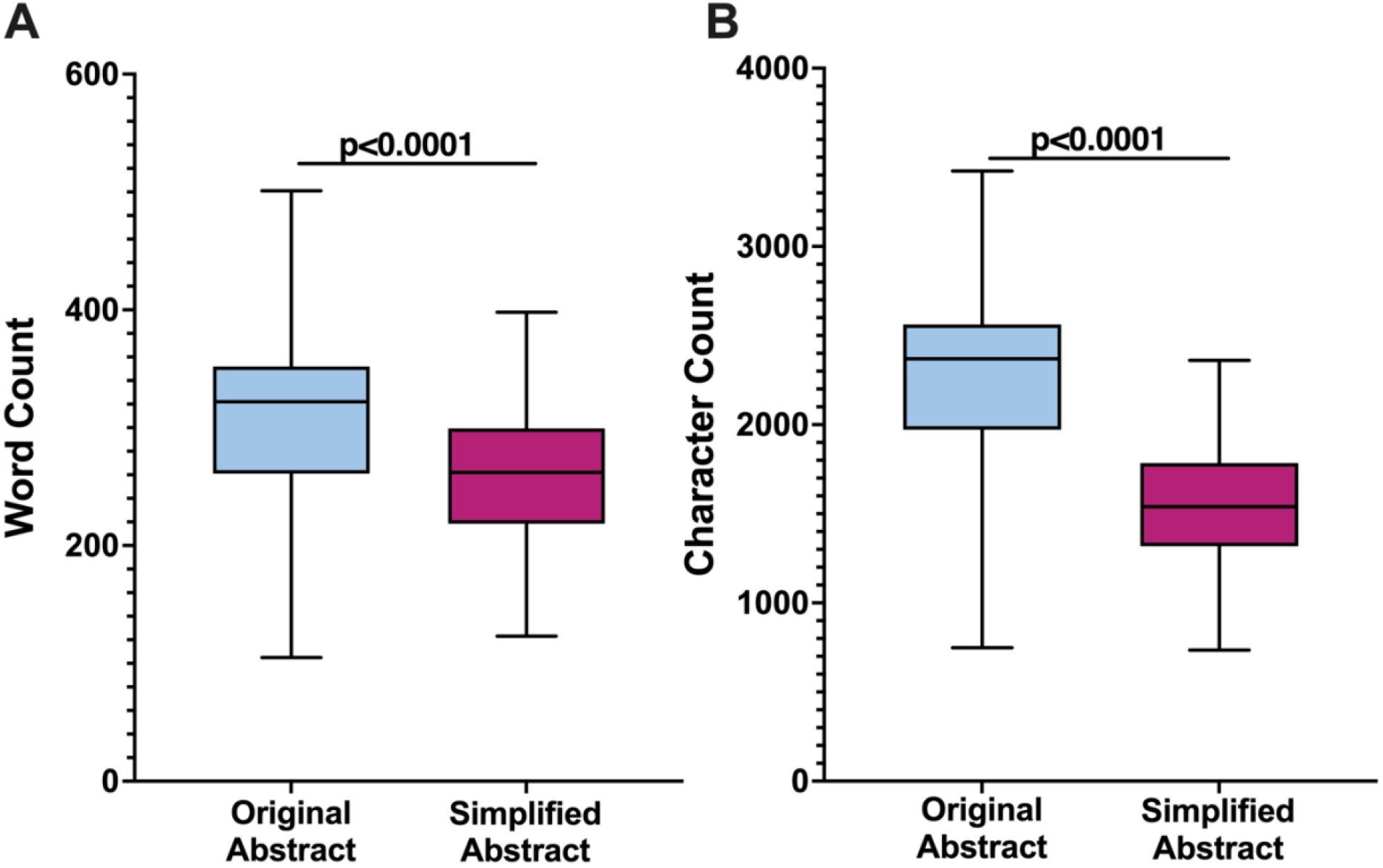
Impact of ChatGPT simplification process on abstract word count and character count. The word count of the abstracts was significantly reduced after simplification (A). Similarly, the character count without spaces was also significantly reduced after simplification (B).

There was also a significant reduction in the educational level required to understand the abstracts, as FKGL average scores decreased from 18.3 (college graduate level) for the original abstracts to 8.6 (8th-9th grade level) for the simplified abstracts; p<0.001 (Fig 2A). FKRE scores increased from 14.6 (college graduate level) to 68.5 (8^th^-9^th^ grade level); p<0.001 (Fig 2B).

**Fig 2.**
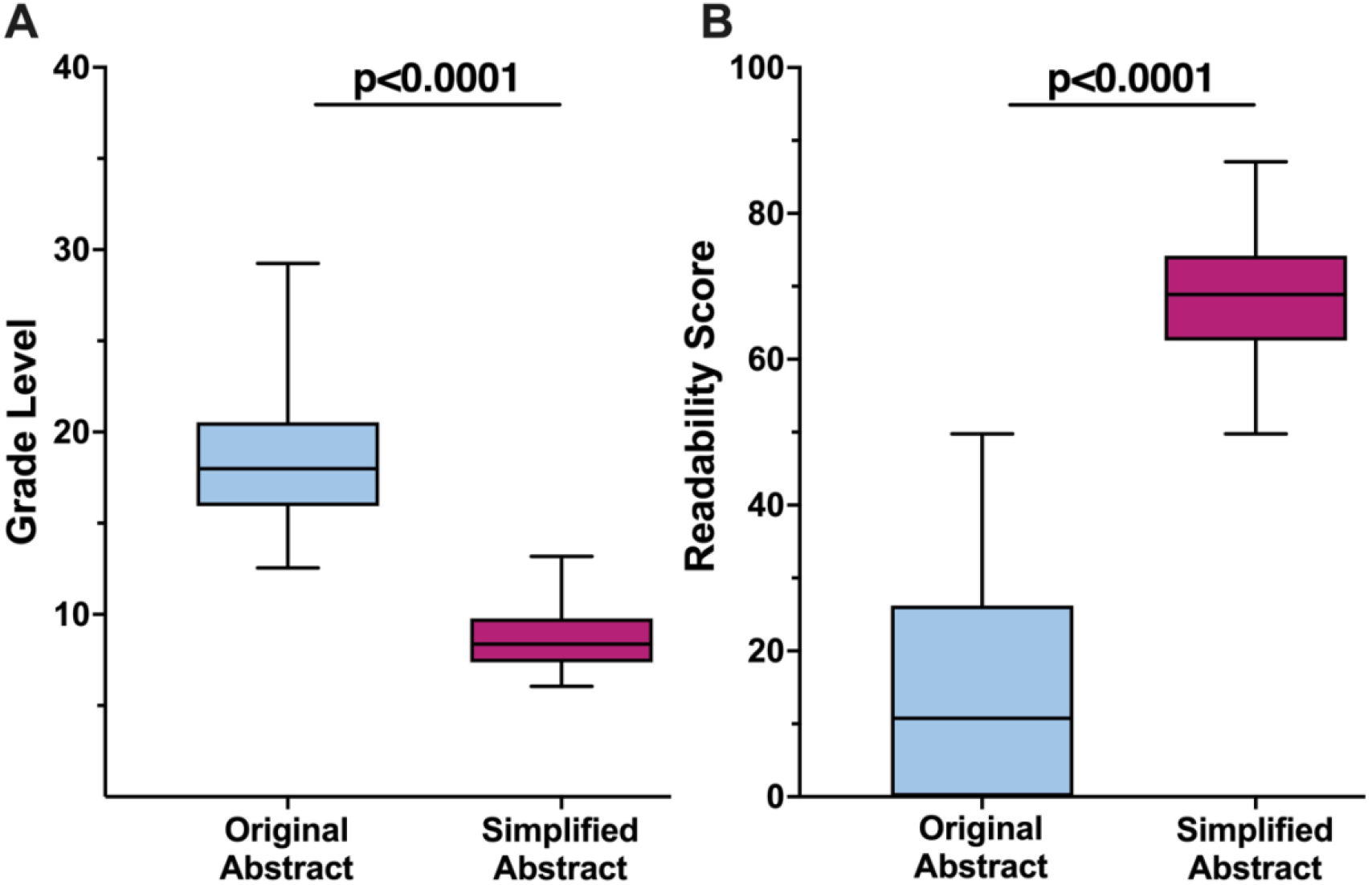
Impact of ChatGPT simplification process on readability. FKGL average scores decreased from 18.3 (college graduate level) to 8.6 (8th-9th grade level after simplification (A). FKRE scores increased from 14.6 (college graduate level) to 68.5, corresponding to an 8th-9th grade level (B). FKGL: Flesch-Kincaid Grade Level; FKRE: Flesch-Kincaid Reading Ease.

Results of the abstract review by physicians and “laypeople” are provided in Tables 1 and 2, respectively. Regarding our physician panel, 76% of the 565 person-abstracts agreed or strongly agreed that no vital content was missing from the simplified abstracts, and 89% agreed or strongly agreed that no incorrect information had been erroneously added to the simplified abstracts. In addition, 94% agreed or strongly agreed that the simplified abstracts were easier to understand, with 91% believing that they would be more comprehensible to patients.

**Table 1.** Results from five-membered physician panel reviewing 113 abstracts each.

| The simplified abstract was missing important information contained in the full abstract | N=565 Person-Abstracts |
| --- | --- |
| Strongly disagree | 302 (53.5%) |
| Disagree | 126 (22.3%) |
| Neither agree nor disagree | 70 (12.4%) |
| Agree | 46 (8.1%) |
| Strongly agree | 21 (3.7%) |
| **The simplified abstract added incorrect information that was not contained in the full abstract** |  |
| Strongly disagree | 400 (70.8%) |
| Disagree | 100 (17.7%) |
| Neither agree nor disagree | 29 (5.1%) |
| Agree | 21 (3.7%) |
| Strongly agree | 15 (2.7%) |
| **The simplified abstract is easier to understand than the full abstract** |  |
| Strongly disagree | 2 (0.4%) |
| Disagree | 6 (1.1%) |
| Neither agree nor disagree | 27 (4.8%) |
| Agree | 108 (19.1%) |
| Strongly agree | 422 (74.7%) |
| **Medical journalists would be able to understand the simplified abstract** |  |
| Strongly disagree | 1 (0.2%) |
| Disagree | 4 (0.7%) |
| Neither agree nor disagree | 7 (1.2%) |
| Agree | 67 (11.9%) |
| Strongly agree | 486 (86.0%) |
| **My patients would be able to understand the simplified abstract** |  |
| Strongly disagree | 2 (0.4%) |
| Disagree | 8 (1.4%) |
| Neither agree nor disagree | 41 (7.3%) |
| Agree | 182 (32.2%) |
| Strongly agree | 332 (58.8%) |
| **My patient would be able to understand the original abstract without looking any terms up** |  |
| Strongly disagree | 195 (34.7%) |
| Disagree | 142 (25.1%) |
| Neither agree nor disagree | 94 (16.6%) |
| Agree | 84 (14.9%) |
| Strongly agree | 49 (8.7%) |

**Table 2.** Results from five-membered layperson panel reviewing 113 abstracts each.

|  |  |
| --- | --- |
| **The abstract was easy to understand** | N=565 Person-Abstracts |
| Strongly disagree | 4 (0.7%) |
| Disagree | 15 (2.7%) |
| Neither agree nor disagree | 33 (5.8%) |
| Agree | 144 (25.5%) |
| Strongly agree | 369 (65.3%) |
| **I did not have to look up any terms to understand the abstract** |  |
| Strongly disagree | 0 |
| Disagree | 17 (3.0%) |
| Neither agree nor disagree | 38 (6.7%) |
| Agree | 112 (19.8%) |
| Strongly agree | 398 (70.4%) |
| **I wish more medical abstracts were simplified to this extent** |  |
| Strongly disagree | 26 (4.6%) |
| Disagree | 102 (18.1%) |
| Neither agree nor disagree | 123 (21.7%) |
| Agree | 167 (29.6%) |
| Strongly agree | 147 (26.0%) |
| **The abstract helped me gain a better understanding of medicine/science/medical science** |  |
| Strongly disagree | 21 (3.7%) |
| Disagree | 73 (12.9%) |
| Neither agree nor disagree | 98 (17.3%) |
| Agree | 205 (36.3%) |
| Strongly agree | 158 (29.7%) |
| **The abstract seemed oversimplified and I felt important information might be missing** |  |
| Strongly disagree | 128 (22.7%) |
| Disagree | 149 (26.4%) |
| Neither agree nor disagree | 97 (17.2%) |
| Agree | 125 (22.1%) |
| Strongly agree | 66 (11.7%) |

The layperson panel classified 91% of the 565 simplified person-abstracts as easy to comprehend. They believed that for 90% of the abstracts, no terms needed to be looked up. However, after reading each abstract, only 56% of the time did a panel member express a desire for more abstracts to be simplified in that manner. Additionally, 66% felt that the abstracts enhanced their understanding of medical science, although responses varied widely regarding whether the abstracts seemed too simplified or potentially omitted crucial information.

## Discussion

Our study evaluated ChatGPT’s efficacy in simplifying medical literature for better patient comprehension. The findings revealed that ChatGPT significantly reduced the reading complexity of cardiology research abstracts from a college graduate level to an 8th-9th grade level, as indicated by the Flesch-Kincaid scores. While prompted to simplify the abstracts to a 5th grade reading level, ChatGPT likely was only able to paraphrase to an 8th-9th grade level due to complex medical terminology, conceptual difficulty, and sentence structure, as scientific abstracts tend to contain longer, compound sentences, making simplification more difficult.

The word and character counts of the abstracts also decreased significantly. Both physicians and laypeople found the simplified abstracts more comprehensible and accurate, although some laypeople expressed concerns over potential oversimplification and the omission of crucial information.

Prior studies have highlighted the challenge of making medical information accessible to patients with varying levels of health literacy. For instance, research by Zaretsky et al. demonstrated ChatGPT’s potential to generate discharge summaries quickly, aiding clinicians in saving time while maintaining content quality [5]. Similarly, another study showed that AI could improve the readability of patient-facing content across multiple National Comprehensive Cancer Network Member Institutions without compromising information quality [6]. Our findings add to this body of work by demonstrating that ChatGPT can effectively simplify complex cardiology research abstracts, making them more accessible to non-medical audiences while largely preserving the integrity of the information. However, despite the overall success in simplifying content, instances of oversimplification or inaccuracies were noted. Some examples, pointed out by both physicians and laypeople, include inaccurate discussion of the differences in outcomes between PCI and CABG, changing the term “microbiota” to “tiny living things in our stomachs,” and describing PCI as a “special blood vessel treatment,” illustrating how the nuance of medical terminology can be lost. These errors, though infrequent, highlight the challenges of ensuring both simplicity and accuracy.

The limited existing literature underscores the mixed reception of AI-generated content among patients. A 2017 case study on DeepMind’s collaboration with the Royal Free London NHS Foundation Trust revealed patient skepticism towards AI-generated medical content [7]. A 2023 survey in Europe found that patients’ perception of AI use in radiology was generally positive but only if it was strictly linked to a supervising radiologist [8]. Our study corroborates these findings, showing that while physicians overwhelmingly found the simplified abstracts accurate and complete, laypeople’s responses were more varied. This suggests a need for further refinement of AI tools to balance simplicity with accuracy and ensure patient trust and acceptance.

Our study had several limitations. The sample size of physicians and laypeople was small and not representative of a broader patient population. Future studies should include a more diverse group with varying education levels, as our panel only consisted of individuals with undergraduate degrees. Additionally, we only used studies from one journal and employed a single generalized prompt. In practice, prompts could be tailored to each patient’s specific health literacy for better personalization.

In conclusion, the integration of AI tools like ChatGPT in healthcare holds significant promise for enhancing patient education by making complex medical information more accessible. Our study indicates that ChatGPT can effectively simplify cardiology research abstracts, improving readability for both medical professionals and laypeople. However, ensuring the accuracy and completeness of the simplified content is crucial. Future research should focus on refining AI capabilities, involving patient feedback, and integrating AI with human oversight to build reliable and trusted resources for patient education.

## Data Availability

Data is available upon request.

## Sources of Funding

No sources of funding

## Conflict of Interest

none declared

## Notes

### Competing Interest Statement

The authors have declared no competing interest.

### Funding Statement

The author(s) received no specific funding for this work.

## References

1. Lopez-Jimenez F, Attia Z, Arruda-Olson AM, Carter R, Chareonthaitawee P, Jouni H, et al. Artificial Intelligence in Cardiology: Present and Future. Mayo Clin Proc. 2020;95:1015-1039. pmid: 32370835

2. Liu W, Laranjo L, Klimis H, Chiang J, Yue J, Marschner S, et al. Machine-learning versus traditional approaches for atherosclerotic cardiovascular risk prognostication in primary prevention cohorts: a systematic review and meta-analysis. Eur Heart J Qual Care Clin Outcomes. 2023;9:310-322. pmid: 36869800

3. Harskamp RE, De Clercq L. Performance of ChatGPT as an AI-assisted decision support tool in medicine: a proof-of-concept study for interpreting symptoms and management of common cardiac conditions (AMSTELHEART-2). Acta Cardiol. 2024;79:358-366. pmid: 38348835

4. Kung TH, Cheatham M, Medenilla A, Sillos C, De Leon L, Elepaño C, et al. Performance of ChatGPT on USMLE: Potential for AI-assisted medical education using large language models. PLOS Digit Health. 2023;2:e0000198. pmid: 36812645

5. Zaretsky J, Kim JM, Baskharoun S, Zhao Y, Austrian J, Aphinyanaphongs Y, et al. Generative Artificial Intelligence to Transform Inpatient Discharge Summaries to Patient-Friendly Language and Format. JAMA Netw Open. 2024;7:e240357. pmid: 38466307

6. Abreu AA, Murimwa GZ, Farah E, Stewart JW, Zhang L, Rodriguez J, et al. Enhancing Readability of Online Patient-Facing Content: The Role of AI Chatbots in Improving Cancer Information Accessibility. J Natl Compr Canc Netw. 2024;22:e237334. pmid: 38749478

7. Powles J, Hodson H. Google DeepMind and healthcare in an age of algorithms. Health Technol (Berl). 2017;7:351-367. pmid: 29308344

8. Ibba S, Tancredi C, Fantesini A, Cellina M, Presta R, Montanari R, et al. How do patients perceive the AI-radiologists interaction? Results of a survey on 2119 responders. Eur J Radiol. 2023;165:110917. pmid: 37327548

